# Association of Sickle Cell Trait with Risk and Mortality of COVID-19: Results from the UK Biobank

**DOI:** 10.1101/2020.07.02.20145359

**Authors:** W. Kyle Resurreccion, Zhuqing Shi, Jun Wei, Rong Na, S. Lilly Zheng, Joseph Hulsizer, Clay Struve, Brian T. Helfand, Janardan Khandekar, Michael S. Caplan, Jianfeng Xu

**Author notes:** please contact the corresponding author, Jianfeng Xu, MD, Dr.PH, at.

## Abstract

We tested the hypothesis that patients with sickle cell trait (SCT), a common condition in individuals of African descent, have increased risk and mortality for coronavirus disease (COVID-19) in the UK Biobank. By June 17, 2020, 1,550 of 7,668 (20%) tested subjects were positive for COVID-19, including 298 (19%) deaths. Blacks had higher rates than Whites for COVID-19 infections (79/222=36% vs. 1,342/7,010=19%, *P*=1.28×10^−9^). Among Blacks, SCT carriers did not have higher infection rates (5/15=33%) than non-SCT carriers (74/207=36%), *P*=1.00. However, SCT carriers had a trend of higher death rates (2/5=40%) than non-SCT carriers (12/74=16%), although not statistically significant (*P* =0.21).

## Introduction

The coronavirus (COVID-19) pandemic has impacted communities worldwide, but its effects are disproportionately expressed in certain groups. The Center for Disease Control reported that non-Hispanic Blacks are approximately five times more likely to test positive for COVID-19 compared to Whites.[1] This finding is supported by UK Biobank (UKB) studies, which found significantly higher positive rates among Blacks even after adjusting for biological (e.g., age, sex, body-mass index, history and number of chronic conditions such as diabetes, asthma, and myocardial infarction), behavioral (e.g., smoking habits, alcohol consumption) and socioeconomic (e.g., education level, employment status, socioeconomic deprivation) factors.[2,3] Additionally, evidence suggests that individuals with preexisting conditions, such as chronic obstructive pulmonary disease (COPD), serious heart conditions, type 2 diabetes, non-allergic asthma, and sickle cell disease (SCD) are particularly vulnerable to COVID-19.[1,4]

The intersection of race and SCD makes it particularly important to consider the impact of COVID-19 in those afflicted. SCD is a genetic disorder that mostly affects people of sub-Saharan African descent, and thus, is common in many Black communities worldwide.[5,6] It is the most common cause of inherited anemia, affecting 80,000-100,000 and 12,500-15,000 individuals in the US and the UK, respectively.[6,7] The disease manifests in individuals who inherit two pathogenic alleles of the gene that codes for hemoglobin (Hgb). While various disease genotypes exist, the manifested phenotype is defective Hgb that makes patients prone to acute episodes of severe pain called vaso-occlusive crises that can lead to fatal acute chest syndrome.[6-9] Individuals with only one pathogenic allele are carriers for sickle cell trait (SCT).[5,6] There are many more carriers than there are SCD patients, and while most are virtually asymptomatic, about 40% of the Hgb in carriers are defective.[5,6]

Because of the novelty of the COVID-19 pandemic, it is yet unknown how individuals with SCT are uniquely affected by the virus due to defective Hgb. The larger number of SCT carriers compared to SCD patients makes the study of this phenotype more relevant to a larger population and more feasible (increased statistical power from larger sample sizes). As such, the goal of this study is to examine the association of COVID-19 and SCT in carriers. Specifically, we hypothesize that: 1) SCT is associated with increased COVID-19 susceptibility, 2) SCT is associated with increased severity of COVID-19, and 3) genetic factors as well as other risk factors and comorbidities significantly modify the risk and severity of COVID-19 in SCT carriers. We test this hypothesis in subjects enrolled in the UKB.

## Methods

This study was performed in the UKB, a population-based study with extensive genetic and phenotypic data for approximately 500,000 individuals from across the UK aged 40-69 at recruitment.[10] Extensive phenotypic and health-related information is available for each participant, including questionnaire, biological measurements, lifestyle indicators, and biomarkers in blood and urine. Follow-up information is provided by linking health and medical records. SCT status, related comorbidities, and risk factors (BMI, diabetes, COPD, hypertension, cancer, stroke and smoking status) were obtained through the International Classification of Diseases-10 (ICD-10) codes and self-report.

COVID-19 test results for the UKB participants are provided by Public Health England (for participants resident in England). This information is updated weekly. Additional COVID-19 data is also collected through a) general practitioner (primary care) data, b) hospital inpatient data, c) death data, and d) critical care data (for COVID-19 positive patients) and updated monthly.

UK Biobank Axiom SNP array data is available for all subjects, while Whole Exome Sequencing (WES) data is available for ∼10% of subjects. Heterozygous Glu6Val mutation was used to identify additional SCT carriers. Blood type (ABO) were inferred from rs8176746 and rs8176719 genotypes,[11] and APOE alleles were inferred from rs7412 and rs429358 genotypes (ε2=T/T, ε3=C/T and ε4=C/C). HLA alleles were obtained from imputed HLA data in the UKB.

Different rates of categorical variables between two groups were tested using the Chi-square test or the Fisher’s exact test when the numbers are small (expected frequencies <5).

## Results and Discussion

By June 17, 2020, COVID-19 test results were available for 7,668 subjects, approximately 1.5% of UKB subjects. Among those tested for COVID-19, 1,550 (20%) were positive and 298 died of COVID-19. Blacks had significantly higher positive rates than Whites (79/222=36% vs. 1,342/7,010=19%, *P* =1.28⨯10^−9^), and higher, but not statistically significant, rates for COVID-19 deaths (14/79=18% vs. 174/1,342=13%, *P* =0.23) (**Table 1**). The mean age at COVID-19 diagnosis was significantly younger in Blacks (63.72) than Whites (68.67), *P* =2.95⨯10^−6^. Similarly, the mean age at COVID-19 death was significantly younger in Blacks (68.93) than Whites (74.39), *P* =1.95⨯10^−3^. The higher positive/death rates for COVID-19 in Blacks than Whites are consistent with other published studies.[2,3] However, to our knowledge, the younger mean age for Blacks who test positive/die from COVID-19 is a novel finding.

**Table 1.** Risk and death of COVID-19 by race and sickle cell trait status in UKB (Based on data from June 14, 2020)

|  |  |  |  |  |  |  |  |  |  |
| --- | --- | --- | --- | --- | --- | --- | --- | --- | --- |
| **Group** | **Tested for COVID-19** |  |  | **Positive for COVID-19** |  |  | **Death due to COVID-19** |  |  |
|  | **No. tested** | **Age, mean (IQR)** | **No. (%) Female** | **No. (%)** | **Age, mean (IQR)** | **No. (%) Female** | **No. (%)** | **Age, mean (IQR)** | **No. (%) Female** |
| White, all | 7,010 | 69.48 (62.48-76.66) | 3,550 (51%) | 1,342 (19%) | 68.67 (59.99-76.74) | 645 (48%) | 174 (13%) | 74.39 (71.43-78.80) | 54 (31%) |
| Black, all | 222 | 64.98 (57.45-73.16) | 131 (59%) | 79 (36%) | 63.72 (56.43-70.03) | 38 (48%) | 14 (18%) | 68.93 (63.77-75.43) | 9 (43%) |
| Black, non-SCT | 207 | 64.96 (57.40-72.73) | 118 (57%) | 74 (36%) | 63.70 (56.50-70.22) | 35 (42%) | 12 (16%) | 68.41 (62.49-74.05) | 4 (33%) |
| Black, SCT | 15 | 65.32 (59.00-74.91) | 13 (87%) | 5 (33%) | 64.00 (56.41-68.55) | 3 (60%) | 2 (40%) | 72.05 (69.79-74.30) | 2 (100%) |

In the UKB, 519 and 166 subjects, respectively, were SCT carriers (ICD-10: D57.3 or heterozygous Glu6Val carriers) and SCD patients (D57.0, D57.1, D57.2, D57.8). The SCT to SCD ratio (519/166=3.13) in the UKB was considerably lower than reported in neonates globally (5,470,000/312,000=17.55),[5] likely due to under-reporting of SCT in ICD diagnosis of UKB. This is supported by the finding that 112 (71%) of the 158 heterozygous Glu6Val carriers detected by WES data did not have ICD diagnosis. Most SCT carriers, as expected, were Black (83%). The proportion of females (70%), however, was significantly higher than the 50% expected for this autosomal inherited trait, *P* =2.00×10^−10^. This observation was found in both Blacks (69%, *P* =5.42⨯10^−8^) and other races (73%, *P* =0.003) and is likely due to a combination of under-reporting of SCT and more women being tested for SCT (e.g., genetic testing for pregnancy).

In Blacks, 15 SCT carriers were tested for COVID-19; 5 were positive and 2 died of COVID-19. In comparison, 207 non-SCT carriers were tested for COVID-19; 75 were positive and 12 died of COVID-19 (**Table 1**). Therefore, SCT carriers did not have higher infection rates (5/15=33%) than non-SCT carriers (74/207=36%), *P* =1.00. However, SCT carriers had a trend of higher death rates (2/5=40%) than non-SCT carriers (12/74=16%), although not statistically significant (*P* =0.21).

The demographic, genetic and clinical characteristics of 15 Black SCT carriers who tested for COVID-19 are presented in **Table 2**. When examining candidate risk factors for COVID-19,[2,12-15] no statistically significant association was found among Black SCT carriers, in part due to small sample size. However, several observations were documented. The mean age of testing was younger in the five COVIID-19 positives (64.18) than the10 negatives (66.16), *P* =0.71. Among the positives, two were current or previous smokers, one had the APOE ε4 allele, and one had blood Type A. In comparison, among the negatives, none were smokers, six carried the APOE ε4 allele, and three were blood Type A. No SCT carriers had the HLA DQA1_509 allele, regardless of positive or negative status for COVID-19. The lower frequency of APOE ε4 allele among COVID-19 positives (33%) than negatives (67%) within this SCT carrier cohort differs from a published study where individuals homozygous for the APOE ε4 allele were significantly more likely to be COVID-19 positive.[13] The lack of a trend between ABO types and COVID-19 risk also differs from published findings that type A and type O subjects have higher and lower COVID-19 risk, respectively.[2,14,15] Finally, the absence of HLA alleles in this cohort also differs from a published study.[2]

**Table 2.** Clinical, demographic, and genetic characteristics of sickle cell trait patients who underwent COVID-19 testing in UK Biobank

|  |  |  |  |  |  |  |  |  |  |  |  |  |  |  |  |
| --- | --- | --- | --- | --- | --- | --- | --- | --- | --- | --- | --- | --- | --- | --- | --- |
| Patient | COVID-19 |  | Demographic and risk factors |  |  |  | Genetic markers |  |  | Comorbidity |  |  |  |  |  |
|  | Diagnosis | Death | Race | Age* | Gender | Smoking | APOE | ABO | HLA DQA1_509 | BMI | Diabetes | COPD | Hypertension | Cancer | Stroke |
| **Black, positive** |  |  |  |  |  |  |  |  |  |  |  |  |  |  |  |
| ID: 4480176 | Yes | Yes | Black | 67.78 | Female | Current | NA | NA | NA | 35.41 | T1D, T2D | N | Y | N | N |
| ID: 5290932 | Yes | Yes | Black | 76.81 | Female | Never | ε3/ε4 | B | 0 | 27.95 | T2D | N | Y | N | N |
| ID: 2322567 | Yes | No | Black | 56.63 | Male | Never | NA | O | 0 | 30.27 | N | N | N | N | N |
| ID: 5828194 | Yes | No | Black | 51.14 | Female | Never | ε2/ε3 | O | 0 | 26.57 | N | Y | Y | N | N |
| ID: 2827022 | Yes | No | Black | 68.55 | Male | Previous | ε3/ε3 | A | 0 | 41.07 | N | N | Y | N | N |
| Summary | --- | --- | --- | 64.18 | 60% | 40% | 33% | --- | --- | 32.25 | 40% | 20% | 80% | 0% | 0% |
| **Black, negative** |  |  |  |  |  |  |  |  |  |  |  |  |  |  |  |
| ID: 1550932 | No | No | Black | 78.29 | Female | Never | ε3/ε4 | B | 0 | 34.46 | T2D | N | Y | N | N |
| ID: 2256003 | No | No | Black | 60.09 | Female | Never | ε3/ε3 | O | 0 | 42.13 | N | N | N | N | N |
| ID: 2557561 | No | No | Black | 58.66 | Female | Never | NA | NA | NA | 28.78 | N | N | Y | N | N |
| ID: 2766417 | No | No | Black | 59.78 | Female | Never | ε3/ε4 | B | 0 | 23.25 | N | N | Y | N | N |
| ID: 2915696 | No | No | Black | 75.07 | Female | Never | ε3/ε4 | B | 0 | 33.31 | N | N | N | N | N |
| ID: 3115703 | No | No | Black | 75.70 | Female | Never | ε2/ε4 | B | 0 | 33.17 | N | N | Y | Multiple myeloma | N |
| ID: 4206423 | No | No | Black | 64.31 | Female | Never | ε3/ε4 | A | 0 | 33.05 | T2D | N | Y | N | N |
| ID: 4900052 | No | No | Black | 53.25 | Female | Never | ε3/ε3 | B | 0 | 30.94 | N | N | N | N | N |
| ID: 3020361 | No | No | Black | 61.38 | Female | NA | ε2/ε4 | A | 0 | 37.23 | N | N | Y | N | N |
| ID: 5839142 | No | No | Black | 75.04 | Female | Never | ε3/ε3 | A | 0 | NA | N | Y | Y | Caecum | N |
| Summary | --- | --- | --- | 66.16 | 100% | 0% | 67% | --- | --- | 32.92 | 20% | 10% | 70% | 20% | 0% |

\* Based on the date of 2020-06-30; NA=Not Available

For comorbidities in Black SCT carriers (Table 2), COVID-19 positives had slightly lower mean BMI (32.25) than negatives (32.92), P=0.83. A higher rate of diabetes was observed in positives (40%) than negatives (20%). It is noted that both patients who died of COVID-19 had diabetes (one had both Type I and II diabetes and another had Type II diabetes only). Modestly higher rates among positives than negatives were also observed for COPD (20% vs. 10%) and hypertension (80% vs. 70%). None of the positives reported cancer. In comparison, two of the 10 negatives had cancer (multiple myeloma and caecum). Stroke was not reported in any Black SCT carriers, regardless of positive or negative status for COVID-19.

One limitation of this study is the relatively small sample size. However, this study represents one of the largest available compared to other published papers and case series that study the association between COVID-19 and SCD/SCT.[7-9] Another limitation is the lack of detailed clinical information on COVID-19 symptoms and treatment. This limits our ability to perform a more comprehensive analysis. Nevertheless, due to the novel and emergent status of this pandemic, more data will likely become available for additional analysis in future studies to address both limitations of this study.

In conclusion, in this first association test between SCT and COVID-19 from a population-based cohort, we find Blacks had significantly higher rates than Whites for COVID-19 infections. The age at COVID-19 infection and death in Blacks were also significantly younger than Whites. Among Blacks, SCT carriers had similar COVID-19 infection rates but a trend of higher, albeit not statistically significant, death rate than non-SCT carriers. Independent larger/more diverse datasets are required to confirm these findings and to expand our knowledge of the association between SCT carriers with COVID-19 susceptibility and severity.

## Data Availability

All datasets used in this study are accessible through the UKB. For more information about data availability, please contact the corresponding author, Jianfeng Xu, MD, Dr.PH, at.

## Additional Information

The UKB was approved by North West – Haydock Research Ethics Committee (REC reference: 16/NW/0274; IRAS project ID: 200778). UKB data was accessed through a Material Transfer Agreement under Application Reference Number: 50295. This study was performed in accordance with the Declaration of Helsinki. All UKB participants gave their informed consent before any data/samples were collected.

All datasets used in this study are accessible through the UKB. For more information about data availability,

## Acknowledgements

We are grateful to the Ellrodt-Schweighauser, Chez and Melman families for establishing Endowed Chairs of Cancer Genomic Research and Personalized Prostate Cancer Care at NorthShore University HealthSystem in support of Dr. Xu and Dr. Helfand, the Rob Brooks Fund for Personalized Prostate Cancer Care at NorthShore University HealthSystem.

## Conflict of Interests

All authors have completed the ICMJE uniform disclosure form at www.icmje.org/coi_disclosure.pdf and declare: no support from any organization for the submitted work; no financial relationships with any organizations that might have an interest in the submitted work in the previous three years; no other relationships or activities that could appear to have influenced the submitted work.

## Author Contributions

Concept: Caplan, Xu

Data analysis: Shi, Wei, Na, Resurreccion

Manuscript draft: Resurreccion, Xu, Caplan

Manuscript revision: All authors

## Notes

### Competing Interest Statement

The authors have declared no competing interest.

### Author Declarations

The UKB was approved by North West - Haydock Research Ethics Committee (REC reference: 16/NW/0274; IRAS project ID: 200778). UKB data was accessed through a Material Transfer Agreement under Application Reference Number: 50295. This study was performed in accordance with the Declaration of Helsinki. All UKB participants gave their informed consent before any data/samples were collected.

